# The Role of Frailty in Shaping Social Contact Patterns in Belgium, 2022-2023

**DOI:** 10.1101/2024.10.10.24315233

**Authors:** Neilshan Loedy, Lisa Hermans, Maikel Bosschaert, Andrea Torneri, Niel Hens

## Abstract

Social contact data are essential for understanding the spread of respiratory infectious diseases and designing effective prevention strategies. However, many studies often overlook the heterogeneity in mixing patterns among older age groups and individual frailty levels, assuming homogeneity across these sub-populations. This oversight may undermine non-pharmaceutical interventions by not targeting specific contact behaviours, potentially reducing their effectiveness in controlling disease. To address this gap, we conducted a contact survey in Flanders, Belgium (June 2022 to June 2023). Using this dataset, we reconstructed frailty-dependent contact matrices and developed a contact-based mathematical model that integrates frailty levels to investigate their impact on disease transmission dynamics. We collected data from 5,723 participants who recorded 31,375 contacts with distinct individuals. Contact patterns were observed to vary based on participants’ age and frailty levels, influenced by the locations of their interactions. Incorporating frailty levels into the mathematical model substantially alters the shape of epidemic curves and peak incidences. Such insights are crucial for designing tailored non-pharmaceutical interventions, indicating the need for similar data collection in different countries.

## 1. Introduction

Individuals within a population exhibit varying characteristics that influence infectious disease transmission [1,2]. These individual-level heterogeneities can arise from various sources, such as varying individual infectiousness or the infectious period (physiological mechanisms) or varying contact rates with infection sources (behavioural mechanisms) [2]. In various environments, individuals’ behavioural heterogeneity can be seen in varying degrees of contact interactions, being dependent on biological ages, genders, health conditions, and social classes (with expected variations in e.g., seasons). These heterogeneities, particularly among older individuals, can significantly impact transmission dynamics. Given that this age group significantly contributes to the infectious disease burden (e.g., 92% of hospitalisations for Herpes Zoster Virus in Italy and 86% for SARS-CoV-2 Virus in England, Scotland, and Wales occur in individuals over 50 years of age [3,4]), understanding contact patterns within this subpopulation is crucial. This need is further amplified by demographic trends suggesting a growing elderly population, which is more susceptible to infections due to age-related immune decline [5,6].

Frail and older populations account for a significant proportion of the health burden, influenced by both biological age and individual health conditions [7]. Therefore, understanding transmission dynamics is crucial for adequately determining suitable public health interventions. However, while transmission models offer valuable insights in understanding disease dynamics, their accuracy is dependent on reliable data on social interactions. This reasoning motivates the collection of social contact data, primarily collected through diary-based social contact surveys, which has been instrumental as an essential tool in parameterising mathematical disease models to understand the dynamics of infectious disease transmission within the population [8-12]. An example is the groundbreaking POLYMOD study, which is a large-scale survey that gathered data from eight European countries with age groups focusing on mainly young adults and adults ^12^. Another example is the CoMix study, which aimed at collecting social contact behavior during the COVID-19 pandemic, highlighting the impact of non-pharmaceutical interventions following the outbreak [9-11,13-18]. However, there are significant knowledge gaps, as social contact surveys that specifically focus on older adults are scarce. This demographic, despite its substantial contribution to disease burden, have been underrepresented in existing studies, hindering comprehensive understanding and precise modeling of disease transmission dynamics within this population.

Although limited, there have been studies on social contacts that specifically target older individuals [19], individuals with chronic illnesses [20], and frail individuals [21]. These studies show that these groups differ not only in the amount of their social interactions but also in the way they interact within the population. Nevertheless, little information is available on which survey and sampling methods are most effective in conducting such a study in this age group. As an illustration, older individuals might encounter challenges when it comes to completing a paper diary, necessitating the need for an in-person interview. It is commonly observed that older individuals tend to favour paper questionnaires as their preferred format, while the suitability of digital approaches for studying health conditions in the older population remains a matter of debate [22]. Since digital questionnaires are faster to distribute (or collect) and pose less burden on a competent participant, exploring the viability of a digital survey holds significance for expanding the survey to other countries, especially in years to come.

This study presents the findings of the Epicurus contact survey study, conducted in Flanders, Belgium. The study explores social mixing patterns across age groups and frailty levels, focusing on older individuals, by including those with chronic conditions and/or residing in healthcare facilities. Furthermore, we analyze how mixing patterns and frailty levels of participants and their contacts might influence disease transmission. Moreover, the study can also serve as a basis for refining data collection protocols, identifying areas for improvement, and informing the design of similar large-scale, multi-country studies in the future across different European countries.

## 2. Methods

### 2.1. Study design

Information on social contacts was obtained cross-sectionally, with the assistance of a market research company. The survey employed a randomized design within the general population, conducted from June 2022 to June 2023. Participants were enlisted (sampling methods) through a database of the National Registry (n.040/2022), acquired from Statbel, and delivered to the market research company. To encompass individuals residing in care facilities, sampling was done through a list of government-recognised facilities. In this study, we refer to a facility which provides long-term housing and a range of services for individuals aged 65 years and older who can no longer live independently at home. These services include household assistance, daily task support, and health care, including nursing care. Different survey methods were used: physical paper forms (paper-based), face-to-face interviews (paper-based), online submissions (Computer-Assisted Web Interface (CAWI)), or through a dedicated app (app-based).

Five target populations were recruited using a stratified sampling approach adjusted by age to ensure a representative sample of the Flanders population (Table S1). The invitation to participate in the survey was divided into three waves: the first wave was conducted during the Summer period (June 14, 2022 - August 18, 2022, and May 10-22, 2023; 20% send-outs), the second wave was carried out in the Fall period (September 5 - December 16, 2022; 40% send outs), and the third wave was conducted in the Winter period (January 23 - April 14, 2023; 40% send outs). This percentage resulted from the intention to include more participants who experience ILI (Influenza-Like Illness) or RSV symptoms during the fall and winter seasons. To achieve the targeted participation rate among individuals aged 22-99 with chronic conditions or experiencing ILI symptoms, additional invitations were sent during fall and winter. These invitations were adjusted by age and mainly sent during winter months to increase the sample size of participants experiencing ILI symptoms. Respondents from these additional invitations who did not have chronic conditions, nor were experiencing ILI symptoms, were categorised into the general population. Sampling in care facilities and the app-based group was terminated upon reaching the desired sample sizes due to cost considerations.

The survey collected demographic information (age, gender, etc.) and vaccination status against influenza and COVID-19. Additionally, participants were asked to complete a contact diary, recording all face-to-face interactions on a specific day (defined as in-person conversations consisting of three or more words, with or without skin-to-skin contact, between 5 am the previous day and 5 am on the survey day), with the number of potentially recorded contacts in a day was limited to 30 (versus 29 to 90 in other studies [12,23]). Details captured for each contact included gender, age range, location(s), intimacy level, frequency, and duration. Participants’ Frailty Index (FI) was measured based on their responses to EuroQol-5 Dimension (EQ-5D) and Short Form Survey-36 (SF-36) questions [24-25], evaluated with the accumulation of deficits approach [26-29]. This well-validated FI captures a broad spectrum of medical conditions (e.g., comorbidities, physical function, and mental well-being) and has been widely used in various international studies [30]. As done by Curran et al. (2021), we divided each participant into one of three subgroups based on their frailty score: non-frail (*FI* ≤ 0.08), pre-frail (0.08 < *FI* ≤ 0.25), or frail (*FI* > 0.25) [29]. If a participant had over 10 quality of life components that were not answered, their Frailty Index (FI) was considered as ‘missing’ unless their available data clearly showed that they already had a high enough score to be classified as frail. Notably, participants aged two years or younger are assumed to be categorized as non-frail as they were not yet able to complete the EQ-5D and SF-36 questions.

### 2.3. Statistical analysis for the number of reported contacts

We developed right-censored negative binomial generalized linear mixed models to examine the factors influencing the average number of contacts reported inside and outside the participant’s home. In this study, the term “home” refers to the domicile of the participant, encompassing houses, apartments, or healthcare facilities. The models incorporated frailty levels and adjusted for other participant characteristics. We performed variable selection using a random forest analysis and the likelihood ratio test (LRT). The reported contacts were right censored at 30 contacts due to a limited number of possible diary entries and were fitted using penalized maximum likelihood within the *‘gamlss’* package in R [31]. Social mixing patterns were further investigated by constructing age-stratified ([0, 50), [50, 60), [60, 750), …, [90, 100)) contact matrices for different locations of contacts (inside or outside the home) [12]. The *‘socialmixr’* package in R was employed with post-stratification weights to account for the distinction between weekdays and weekends when generating the contact matrices [32].

### 2.4 Mathematical compartmental transmission model

In this study, we utilize a discrete-time age-structured compartmental model which incorporates contact matrices for age-specific transmission rates, previously developed by Abrams et al. (2021) [9]. The model is extended to account for age and frailty mixing patterns, enabling the investigation of a COVID-19-like disease’s spread and assessing the impact of frailty-dependent interactions within the Belgian population **(Supplementary Material)**. This model assumes that individuals become infectious (symptomatic or asymptomatic) after a latent period. Symptomatic cases may progress to severe illness requiring hospitalisations or being admitted to the intensive care unit (ICU), where they are assumed to be isolated and can not transmit the disease. We initialized our disease transmission model on March 1st, 2020, to reflect the early stages of the COVID-19 pandemic (Wuhan strain, without vaccination). Simulations were conducted for the Belgian population of 2020 (timestep: 1 day) over 100 days.

The model uses parameter values from previous studies on COVID-19 vaccination strategies, which were calibrated in parallel with this study **(Table S2)** [9,33]. To tailor the model to the observed data, we set the initial population and distribution of infected individuals across age groups and frailty levels to be proportional to the survey data **(Table S3)**. To compare the impact of different mixing assumptions on transmission dynamics, we conducted two analyses: one using a constant proportionality factor (*q*) capturing host- and disease-specific characteristics, and the other using a constant basic reproduction number (*R*_0_ = 2.90) calculated with the next-generation matrix approach for COVID-19 [9,34]. The effects of various contact-level assortativity on transmission dynamics were analyzed using three different scenarios: proportional, uniform, and full assortativity. In the proportional scenario, contact-level assortativity reflects the proportion of frailty levels obtained from the survey. In the uniform scenario, the assortativity is identical across all frailty levels. Lastly, the full assortativity scenario simulates individuals who only come into contact with others who have the same level of frailty. To isolate the effect of altered contact patterns on disease spread, we assumed homogeneous parameters across frailty levels. This simplification enabled us to attribute any observed changes in transmission patterns primarily to variations within the contact matrices.

## 3. Results

### 3.1 Study population

Approximately 31,000 letters (55% main and 45% additional invitations) were dispatched to the participants within the random sample generated by the National Registry. The overall response rate was 19.34% (*n* = 5,995). The majority of subpopulations from the collected sample exceeded the desired target study population, except for the ILI subpopulation, which only reached 0.3% of the expected quota **(Table S1)**. Notably, a preference for the paper-based survey (66.67%) was observed compared to the computer-based version (CAWI or app-based) (33.32%). Of the initial respondents, 272 individuals did not consent to participate, and one participant was excluded because they did not comply with the contact reporting guidelines (the reported contact was not from the day preceding the survey). The final analysis included 5,723 consenting participants (44.18% male, 53.45% female) who reported 31,375 contacts **(Table S4)**.

The mean participant age was 53.4 years, with a median of 59 years (IQR: 37-71). Around 70% and 90% of participants who used the Computer-Assisted Web Interface (CAWI) and app-based approach, respectively, were below the age of 60 years. Conversely, the paper-based survey skewed towards individuals over 60 **(Figure S1)**. The data indicates that individuals over 70 years old generally tend to engage in volunteer work or remain unemployed (either retired or capable of working but not currently employed). Chronic health conditions were reported by 33.4% of participants, all exceeding 50 years old. Most of the participants resided in houses or apartments of size 2 (33.2%) or 3 (18.4%), while a small proportion (3.5%) resided in long-term care facilities (retirement or nursing homes (ROB: *Rustoord voor Bejaarden*; RVT: *Rust-Verzorgingstehuis*)), especially older individuals. Furthermore, frailty assessment revealed a significant association with biological age, with older participants tend to be in the more frail categories. Among the total sample, there were 830 participants (14.50%) classified as frail, 1,749 (30.56%) as pre-frail, 2,681 (46.85%) as non-frail, and 463 (8.09%) were categorized as missing, as described in Section 2.1.

### 3.2 Contact behavior and mixing patterns

On average, participants report 5.48 contacts daily, with a median of 4 contacts (IQR: 2-7) **(Table S5)**. When considering contacts reported outside the household **(Figure 1)**, we observed a notable association between frailty and the number of reported contacts. Frailty status influences the average number of contacts, with frail individuals reporting fewer contacts compared to non-frail and pre-frail individuals. This decrease in contact varies with the degree of frailty and is more pronounced in older age groups. Note that the small number of non-frail individuals in the 80-89 and 90-100 age categories (0.8% and 0.2% of non-frail individuals, respectively) may lead to a noticeable discrepancy between the observed average number of contacts and the model-based number of contacts.

**Figure 1:**
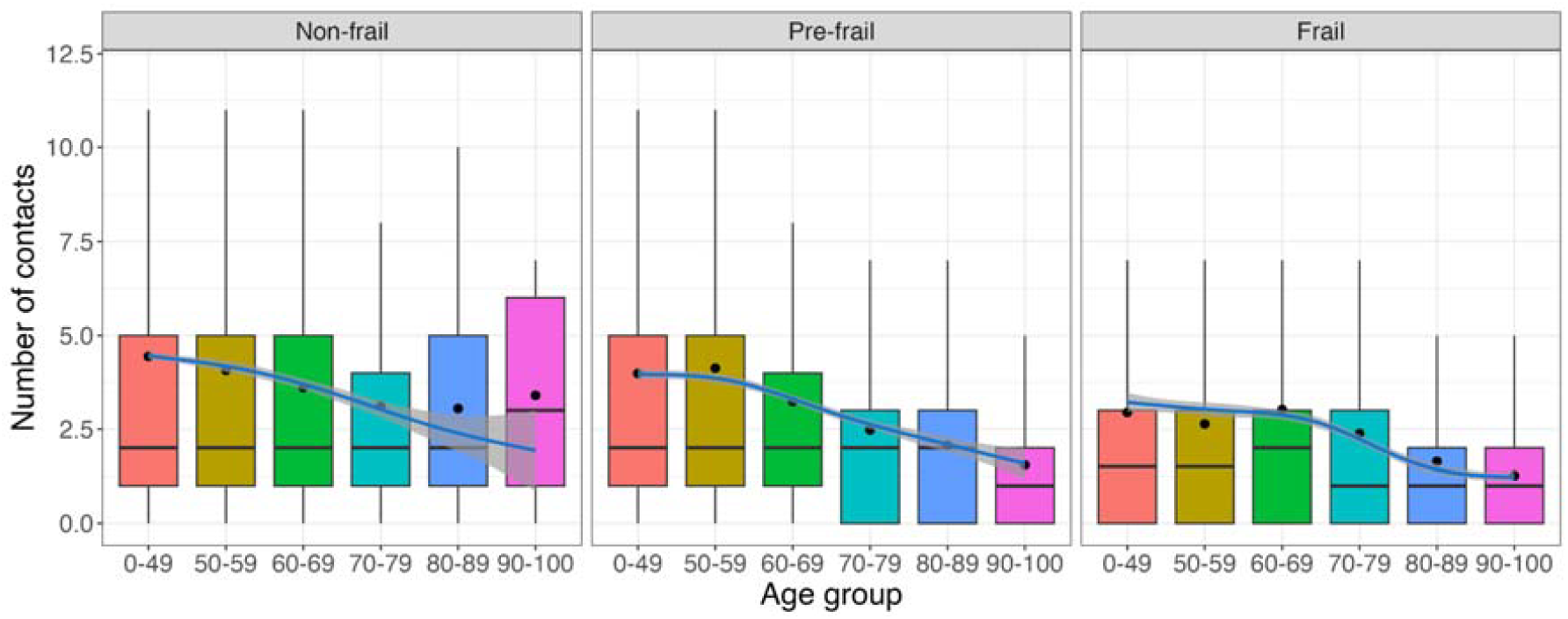
Number of contacts reported outside the household per participants’ age group by their frailty level. Plots show the distribution of contacts, the average number of contacts (dots) and model results (mean as solid lines and 95% confidence intervals as shaded area).

A significant portion of contacts occurs with individuals of similar age, as indicated in the contact matrices for which higher values are reported on the diagonal **(Figure S2)**. More precisely, older individuals (60-90 years) primarily interact with others in their age range, followed by interactions with younger adults (30-59 years). Intergenerational mixing occurred more inside participants’ homes, particularly between younger individuals (under 18) and adults (30-50 years). Higher contact rates are observed when comparing contacts outside participants’ homes with those reported inside. Pre-frail and non-frail individuals reported more contacts outside their homes compared to frail individuals **(Figure 2)**. In contrast, frail individuals tended to report more contacts inside their homes. For individuals under 70 with frailty, these trends persisted with a higher number of outside contacts reported compared to those over 70 with frailty, who tended to have more contacts inside their homes. When considering participants with chronic conditions, we observe higher contact rates outside their homes, except for those over 70 years old, who reported more contacts within their homes **(Figure S3 - Figure S5)**.

**Figure 2:**
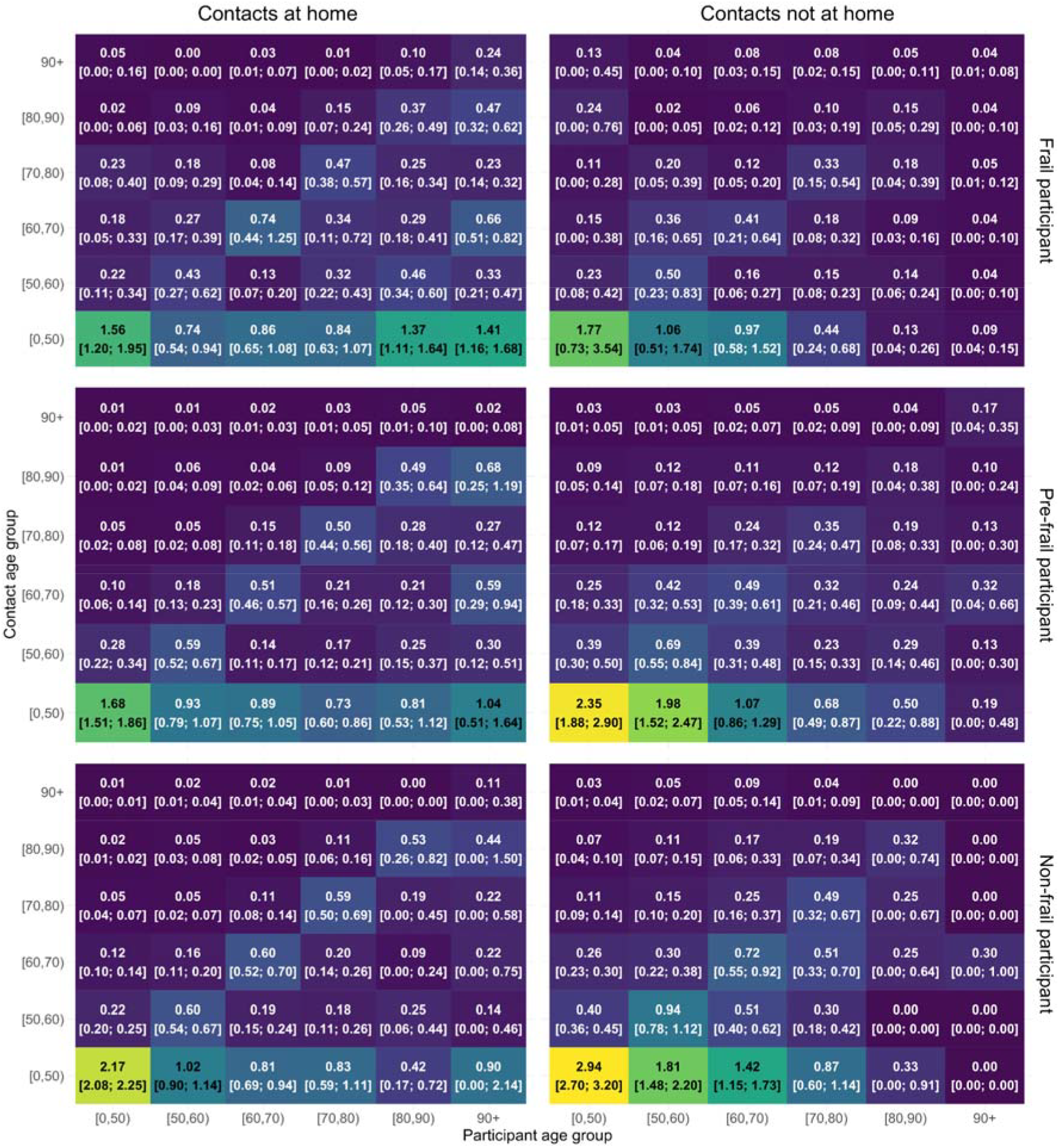
Contact matrices showing the mixing patterns of participants based on their frailty level for contacts reported inside and outside the household, together with 95% confidence intervals obtained from non-parametric bootstrap.

### 3.2 Contact characteristics

Variables expressing the number of contacts are selected based on the results of both random forest analysis **(Figure S6)** and the likelihood ratio test **(Table S6). Table S7** presents the relative incidence (RI) with its 95% confidence interval (CI) obtained from GAMLSS models inferring the number of indoor and outdoor contacts based on these selected covariates. While holding other variables constant, the sample drawn from the care facilities reported more contacts (RI = 2.141 [1.797 - 2.550]) inside the home (**Table S7**). The gender variable had a significant effect on the number of contacts reported, with women reporting a higher number of contacts inside (RI = 1.077 [1.012 - 1.146]) and outside (RI = 1.066 [1.020 - 1.113]) the home. People who completed the questionnaire on paper tend to report more contacts, with a relative incidence of 2.293 [2.136 - 2.463] and 1.160 [1.106 - 1.215] for outside and inside home contacts, respectively. Household size had different effects on inside and outside contacts; participants who lived alone tended to report more outside contacts (RI = 1.171 [1.054 - 1.298]), but this pattern was reversed for inside contacts (RI = 0.811 [0.731 - 0.900]). Unemployed people reported more contacts inside the home (RI = 1.087 [1.001 - 1.180]) compared to outside the home (RI = 0.641 [0.573 - 0.716]), and compared to people who worked full time. Lastly, education also has a significant effect on out-of-home contacts, with participants holding undergraduate degrees generally reporting more contacts. More out-of-home contacts were reported during weekdays and non-holidays. There was an interaction between age and holidays on contacts, with older participants (above 50 years old) reporting fewer contacts during non-holidays, compared to children under nine years old. Non-frail individuals significantly report more contacts outside the home. Lastly, it was also found that there was an interaction between reported contacts and frailty level and whether or not participants completed the survey with assistance.

### 3.3 Mathematical compartmental transmission model

The results of the simulation study indicate varying disease dynamics with a constant value of q across all mixing patterns **(Figure S7)**. Full assortativity results in significantly lower peak epidemics (2.36, 0.45, 0.05 per 100,000 for non-frail, pre-frail, and frail groups, respectively), while uniform mixing yields the highest peak epidemics (5.01, 1.95, 0.74 per 100,000 for non-frail, pre-frail, and frail groups, respectively), compared to other scenarios. When simulating epidemic outbreaks and keeping a constant value of *R*_0_, we observed mixing patterns among different frailty levels affect disease transmission dynamics **(Figure 3)**. While attack rates across the three scenarios - proportional (0.78), uniform (0.81), and fully assortative (0.82) - show only a slight difference, the underlying patterns of disease transmission vary considerably. Under the full assortativity scenario, non-frail and pre-frail individuals experience the fastest epidemic peak (day 28) and highest incidence rates (5.92 and 1.92 per 100,000, respectively). Conversely, frail individuals show the slowest peak (day 30) and lowest incidence rate (0.47 per 100,000). Uniform mixing results in the fastest and highest incidence rate for frail individuals (0.70 per 100,000 after 26 days), while proportionate mixing shows the lowest incidence rate and slowest peak for non-frail individuals (3.96 per 100,000 in 32 days).

**Figure 3:**
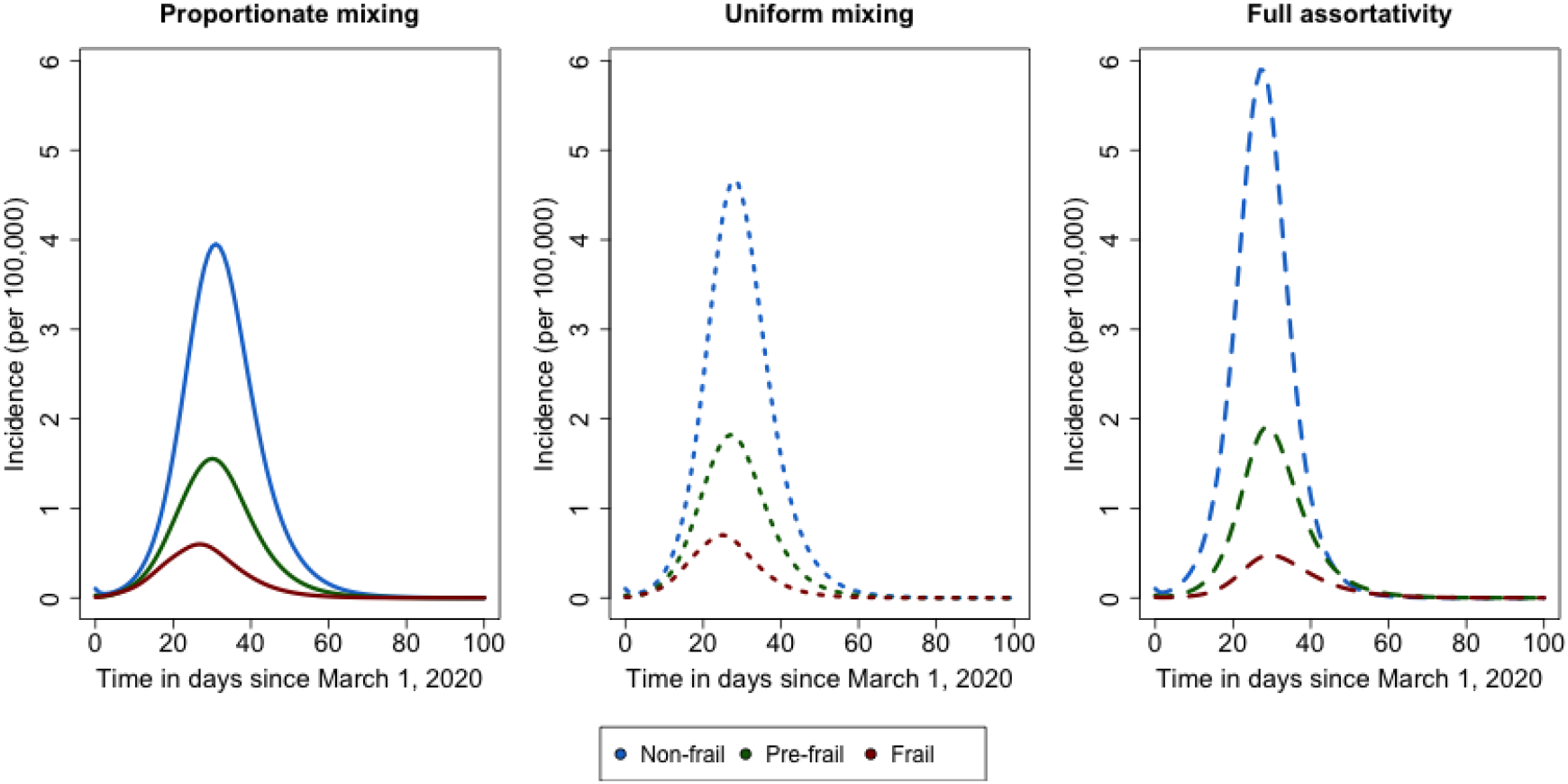
Comparison of COVID-19-like epidemic curves for the Belgian population with various frailty-based mixing patterns.

## 4. Discussion

In this manuscript, we presented the outcomes of the Epicurus study, which was conducted to investigate social mixing behaviors in the Flemish region, Belgium, from June 2022 to June 2023. In particular, we focused on older individuals, as existing literature highlights their significant role in disease transmission [5,20,21], yet there is still a knowledge gap concerning how their behaviors vary with frailty levels. We conducted simulations using a compartmental model to describe infectious disease spread, incorporating frailty-dependent mixing patterns obtained from the study as a proxy of age-specific transmission rates, and compared outbreak characteristics under various degrees of interactions. In particular, we have considered the spread dynamics of SARS-CoV-2, given its recent prominence and significant impact.

The present work gives important insights for the design of future data collection. In particular, we observed a notably high response rate when participants were randomly selected from the national registry. Residents of healthcare facilities demonstrated a keen willingness to participate in the study, facilitating a smooth data collection process. We found that the paper version of the survey was preferred over the computer-assisted web interface (CAWI) or app versions. However, managing incomplete responses posed a challenge as there was no means to validate the provided answers. Recruiting individuals aged 50-75 years from healthcare facilities presented difficulties, as this demographic typically does not reside in such facilities unless they have specific medical conditions. This challenge also extended to individuals with ILI symptoms, as they may not be inclined to participate in surveys when feeling unwell. In future studies, improving data on ILI symptoms could involve asking participants about their symptoms and contacts during their most recent ILI period. This approach may help address the issue, albeit susceptible to recall bias [16]. Alternatively, supplementing national registry data with additional information from general practitioners could be of added value. When doing so, potential bias towards specific subpopulations should be carefully considered.

Our findings indicate distinct contact patterns among different age and frailty groups, with a particular emphasis on the location where contacts took place. Older frail individuals predominantly interacted within their homes, potentially increasing the risk of intra-household outbreaks. This contrasts with younger frail individuals who exhibited more external contacts, suggesting interactions between age, frailty, and mixing patterns. These findings extend previous work by Backer et al. (2023), who reported overall reduced contact rates among frail individuals, by highlighting the disproportionate decrease in external contacts, potentially due to factors such as physical limitations or social isolation [21]. Understanding the long-term implications of these distinct contact patterns for disease transmission is essential for designing effective non-pharmaceutical interventions. Tailoring strategies to limit disease spread to the specific contact behaviors of vulnerable groups, such as older frail individuals, can optimize public health efforts by mitigating healthcare burdens, leading to better health economic outcomes. Additionally, incorporating healthcare costs that depend on frailty into cost-effectiveness calculations within mathematical models can enhance their accuracy, leading to more realistic decision-making outcomes [35].

We further emphasize the impact of considering heterogeneity from varying frailty levels within populations on the spread of respiratory infectious diseases, using a contact-based deterministic mathematical model. While maintaining a constant transmission rate across frailty groups, we observed distinct transmission dynamics. This emphasizes the need for caution when extrapolating population-level parameters to specific subgroups. Furthermore, we investigated the impact of frailty levels on transmission dynamics by maintaining a constant reproductive number. Our results demonstrate significant variations in epidemic outcomes across different frailty groups, driven by distinct contact patterns [36]. The presented mathematical model successfully highlights that reliable disease transmission modeling necessitates a thorough understanding of heterogeneity in mixing patterns, as ignoring these factors can lead to inaccurate predictions, suboptimal intervention strategies, and misguided economic evaluations [37]. Future research can enhance the model’s realism by incorporating variations in susceptibility and infectivity, which are key factors in transmission dynamics, especially when considering the different frailty levels within individuals.

The collected data and analysis performed emphasize the importance of collecting detailed information on contact patterns and investigating their potential role in shaping disease dynamics, particularly within older individuals and those with varying degrees of frailty. In addition to obtaining preliminary insights into the social mixing behavior within this cohort, the study can also be utilized to test the questionnaires and assess the feasibility of conducting a large-scale contact study. However, it is important to note that our scope does not encompass comparisons across countries, including any analysis before, during, or after the COVID-19 pandemic, nor does it delve into future projections. Extrapolating these results to other countries may not be wise, as each country possesses unique characteristics (e.g., cultural and educational background, infrastructure, and social structure) that may considerably impact the validity of such extrapolations [38]. However, the insights and methodologies discussed herein can guide potential extensions of this study to a European context.

Our study considers frailty alongside chronic disease, as it offers a broader overview of individual health levels [39]. Nevertheless, we utilized a singular method to calculate frailty, potentially overlooking variation in contact patterns across different frailty measurements. Potentially, employing a broader frailty index calculation that encompasses multiple health indicators could provide deeper insights into the interplay between frailty and contact behavior. However, constructing such indices can be cumbersome and may induce participant fatigue if the questionnaire becomes overly lengthy [16]. Future investigations could employ multidimensional frailty scales to unveil a deeper understanding of how frailty influences social interactions. Additionally, while COVID-19 and flu vaccinations did not significantly impact the number of contacts in this study, exploring the effects of vaccines for other diseases, such as RSV and pneumococcal infections, may provide valuable insights, particularly for the elderly and high-risk groups [40]. Therefore, future research could delve deeper to explore these aspects.

## 5. Conclusion

The Epicurus study in Flanders, Belgium, aimed to characterize the social contact patterns of older individuals and those with varying levels of frailty in relation to disease transmission. Our findings revealed distinct contact patterns across different frailty levels. By integrating these patterns into contact-based mathematical modeling, we demonstrated the critical importance of accounting for frailty-dependent heterogeneity in disease transmission. These insights contribute to our understanding of how frailty affects contact patterns and disease spread, emphasizing the need for further data collection and analysis across a broader population.

## Ethics declarations

Ethical approval was obtained from the Comite voor Medische Ethiek UHasselt (CME UHasselt) with Reference Number: CME2021/110.

## Data availability statement

The datasets generated during and/or analyzed during the current study are available from the corresponding author upon reasonable request.

## Acknowledgement

The authors gratefully acknowledge the IMI VITAL project for their valuable input and feedback during the development of the study protocol. We extend our sincere thanks to the Ipsos team for conducting the survey, collecting data, and facilitating the rapid progress of this study. We especially appreciate the exceptional project management support provided by Sarah Vercruysse. All important findings will be informed to the IMI VITAL WP3.

## Funding

Funding for this study [study number: 215366] was provided by GSK. GSK was provided the opportunity to review a preliminary version of this publication for factual accuracy, but the authors are solely responsible for final content and interpretation.

## Author Contributions

NH and LH initiated the study. NH and LH designed the initial survey. NL and LH cleaned and prepared the data for the platform. NL did the formal analysis and wrote the original draft with the supervision of LH, MB, AT, and NH. All authors contributed and reviewed the manuscript and approved the final version for publication.

## Competing Interest

The authors have declared that no competing interests exist.

## Notes

### Competing Interest Statement

The authors have declared no competing interest.

