## Supplementary material for "The Role of Frailty in Shaping Social Contact Patterns in Belgium, 2022-2023"

**Table of Contents:**

**1. Decomposition of social contact matrices 1**
**2. Mathematical compartmental transmission model 2**
**3. Supplementary materials (Figures) 4**
**4. Supplementary materials (Tables) 8**

#### 1. Decomposition of social contact matrices

To explore the impact of behavioural changes following the frailty status of individuals, we derive social contact matrices to reflect mixing patterns between people with different frailty statuses. The social contact matrix, denoted as $M,$ represents the average number of daily contacts reported by individuals within specific age classes. Here, each element $m_{ij}$ depicts the mean number of contacts reported by participants in age class $i$ with individuals in age class $j$. In this study, we define three distinct matrices, namely $M_{frail}$, $M_{pre-frail}$ $and$ $M_{non-frail}$, delineating mixing patterns associated with individuals classified as frail, pre-frail and non-frail with the general population, respectively. Let $W_y$ be a matrix with element $w_{y,i\cdot}$ representing the proportion of individuals in an age category $i$ falling into a health condition $y$ ($y =$ frail, pre-frail, non-frail). This can be defined as:

$$W_y = \frac{w_{y,i\cdot}}{\sum_{a=1}^{i} w_{y,a}}.$$

With $w_{y,i\cdot}$ is the total number of individuals with a health condition $y$ in age class $i$ and $\sum_{a=1}^{i} w_{y,a}$ is the total number of individuals in a health condition $y$. Based on the law of total expectation, the conditional expected number of contacts made by individuals can be written as:

$$E(X = M) = \sum_{y=1}^{3} E(X = M \mid Y = y) \cdot W(Y = y).$$

It is important to note that we can decompose the mixing patterns further, by looking at the fact that individuals with a frailty level $y$ can interact with others in any of the frailty categories (e.g., $M_{frail,\, non-frail}$ represents contacts between frail and non-frail individuals). As such, contacts made by individuals in a frailty condition $y$ can be written as:

$$M_y = \sum_{y'=1}^{3} M_{y,y'}.$$

Let $\xi_{y'} \in \mathbb{R}$, such that $0 \leq \xi_{y'} \leq 1$ be the degree of assortativity with individuals in health condition $y'$, with $\sum_{y'=1}^{3} \xi_{y'} = 1$ and $\rho_{i-y,\, j-y'}$ is the matrix of probabilities with each element representing the probability of an individual with the frailty status $y$ in age class $i$ making contact with an individual in age class $j$ with frailty status of $y'$. This can be written as:

$$\rho_{i-y,\, j-y'} = \frac{w_{y',i\cdot}\, \xi_{y'}}{\sum_{y'=1}^{3} w_{y',i\cdot} \xi_{y'}}.$$

Hence, contact matrices that reflect mixing patterns for individuals in the frail status $y$ can be defined as:

$$M_{y,\, y'} = M_y \times \rho_{i-y,\, j-y'}.$$

#### 2. Mathematical compartmental transmission model

The following set of ordinary differential equations describes the flows in the proposed age-structured compartmental model:

$$\frac{d\mathbf{S_y(t)}}{dt} = -\mathbf{S_y(t)}\lambda_\mathbf{y}(\mathbf{t})$$
$$\frac{d\mathbf{E_y(t)}}{dt} = \lambda_\mathbf{y}(\mathbf{t})\mathbf{S_y(t)} - \gamma\mathbf{E_y(t)}$$
$$\frac{d\mathbf{I_{presym,y}(t)}}{dt} = \gamma\mathbf{E_y(t)} - \theta\mathbf{I_{presym,y}(t)}$$
$$\frac{d\mathbf{I_{asym,y}(t)}}{dt} = \theta p\mathbf{I_{presym,y}(t)} - \delta_1\mathbf{I_{asym,y}(t)}$$
$$\frac{d\mathbf{I_{mild,y}(t)}}{dt} = \theta(1-p)\mathbf{I_{presym,y}(t)} - \{\psi + \delta_2\}\mathbf{I_{mild,y}(t)}$$
$$\frac{d\mathbf{I_{sev,y}(t)}}{dt} = \psi\mathbf{I_{mild,y}(t)} - \omega\mathbf{I_{sev,y}(t)}$$
$$\frac{d\mathbf{I_{hosp,y}(t)}}{dt} = \phi_1\omega\mathbf{I_{sev,y}(t)} - \{\delta_3 + \tau_1\}\mathbf{I_{hosp,y}(t)}$$
$$\frac{d\mathbf{I_{icu,y}(t)}}{dt} = (1-\phi_1)\omega\mathbf{I_{sev,y}(t)} - \{\delta_4 + \tau_2\}\mathbf{I_{icu,y}(t)}$$
$$\frac{d\mathbf{D_y(t)}}{dt} = \tau_1\mathbf{I_{hosp,y}(t)} + \tau_2\mathbf{I_{icu,y}(t)}$$
$$\frac{d\mathbf{R_y(t)}}{dt} = \delta_1\mathbf{I_{asym,y}(t)} + \delta_2\mathbf{I_{mild,y}(t)} + \delta_3\mathbf{I_{hosp,y}(t)} + \delta_4\mathbf{I_{icu,y}(t)}$$

We initialised our disease transmission model on March 1st, 2020, reflecting the early stages of the COVID-19 pandemic when no vaccines were yet available and only the original strain of the virus was circulating. Let $y = 1, 2, 3$ be the frailty status representing frail, pre-frail, and non-frail individuals and age-specific force of infection in age group $k = 1, 2, ..., K$ with frailty status $y$ is denoted by $\lambda_y(k, t)$. This force of infection represents the instantaneous rate at which a susceptible person in age group $k$ with frailty status $y$ acquires infection at time $t$. As such, the force of infection is defined as

$$\lambda_y(k, t) = \sum_{y'=1}^{y} \sum_{k'=1}^{k} \beta_{y,y'}(k, k') I_{y'}(k', t).$$

Let $c_{y,y'}(k, k')$ are the per capita rates at which an individual with frailty status $y$ in age group $k$ makes contact with an individual with frailty status $y'$ in age group $k'$, per unit of time, and $q$ is a proportionality factor capturing contextual and host- and disease-specific characteristics such as susceptibility and infectiousness. Relying on the so-called social contact hypothesis, we have

$$\beta_{y,y'}(k, k') = q \cdot c_{y,y'}(k, k'),$$

which can be defined as the transmission rates for individuals in an age group $k$ that make contact with an individual in an age group $k'$, per unit of time and $I_{y'}(k', t)$ denotes the total number of infectious individuals in an age group $k'$ with frailty status $y'$ at time $t$.

### 3. Supplementary materials (Figures)

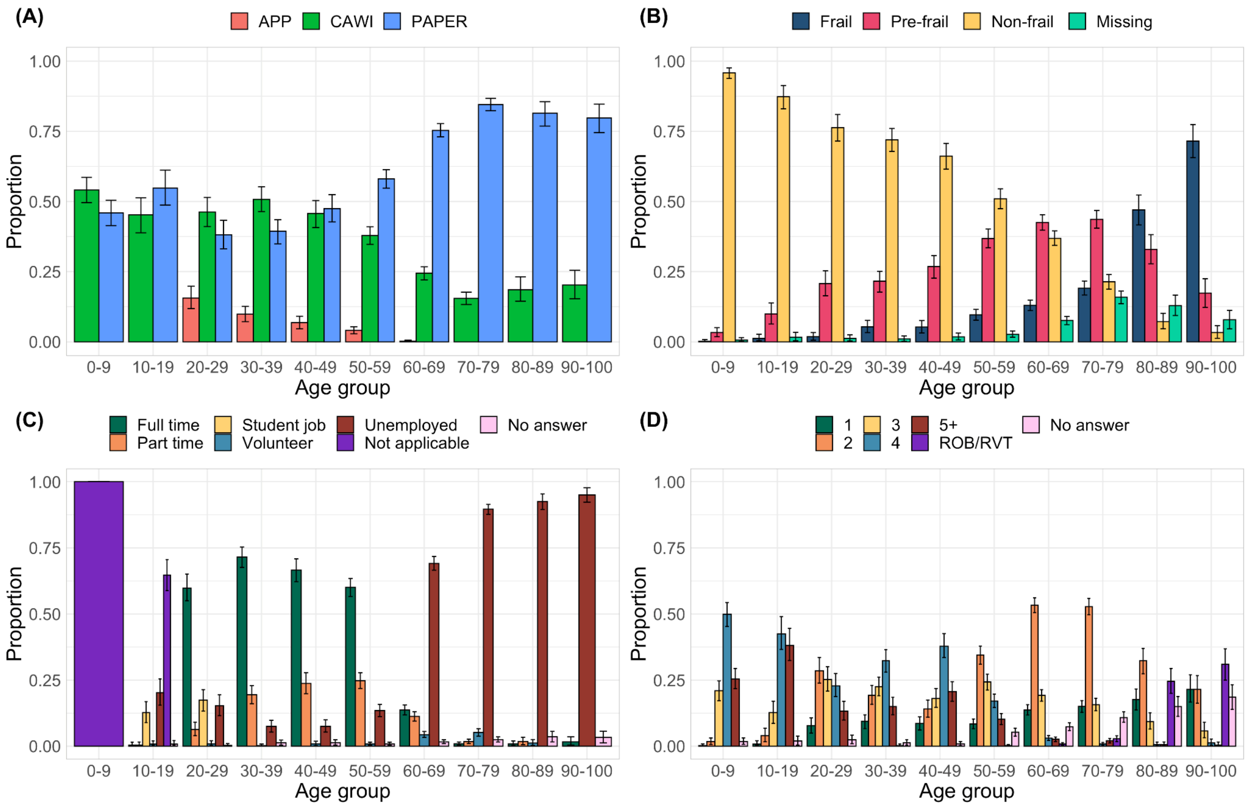


**Figure S1:** Participant characteristics of the study population. **(A)** Fractions of participants with their corresponding survey methods by age group **(B)** Fractions of participants by frailty level **(C)** Fraction of participants by occupancy **(D)** Fraction of participants with household size.

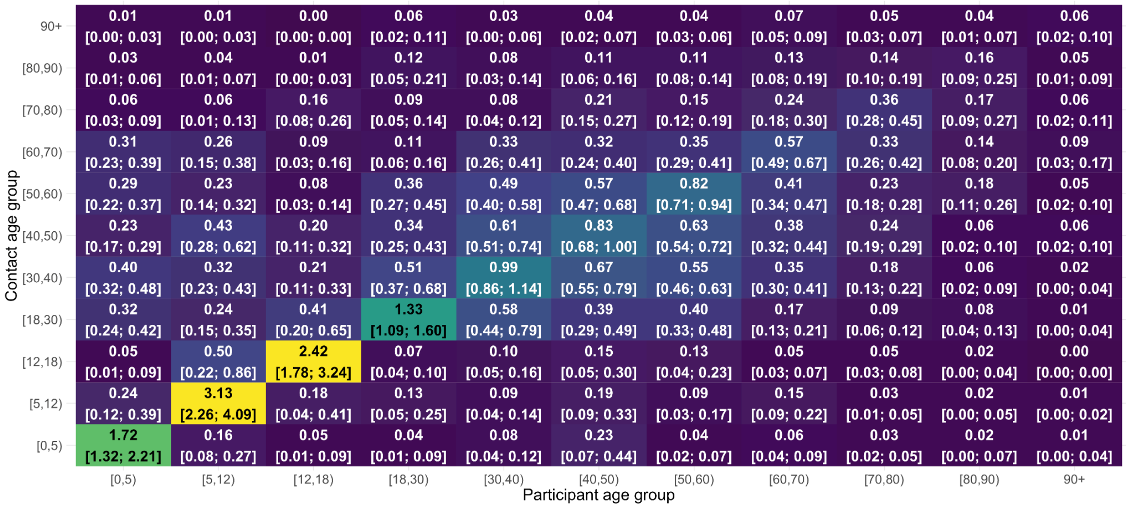

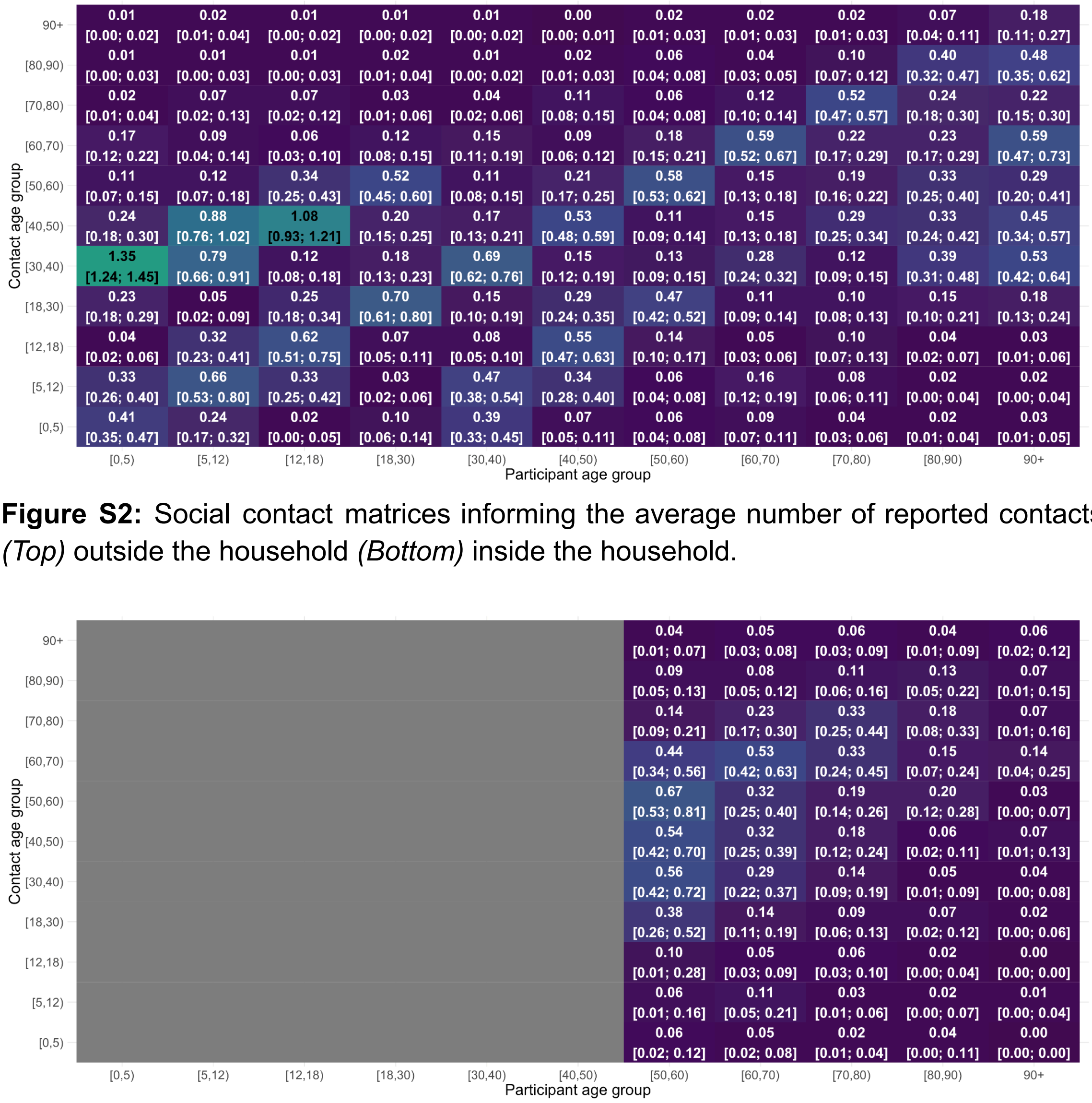


**Figure S2:** Social contact matrices informing the average number of reported contacts *(Top)* outside the household *(Bottom)* inside the household.

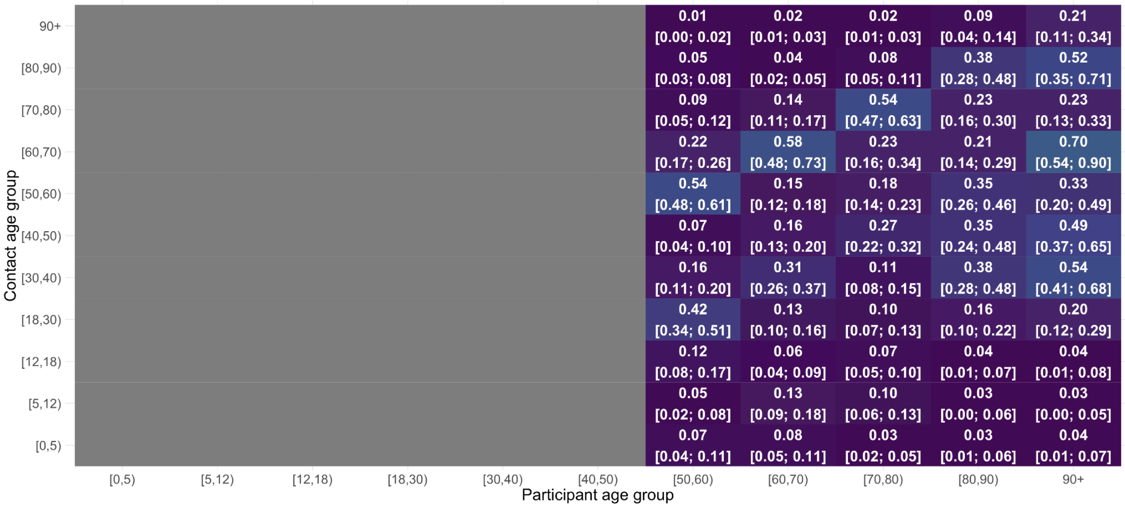


**Figure S3:** Social contact matrices for chronic participants *(Top)* outside the household *(Bottom)* inside the household. Note that only participants older than 50 years old reported having a chronic condition.

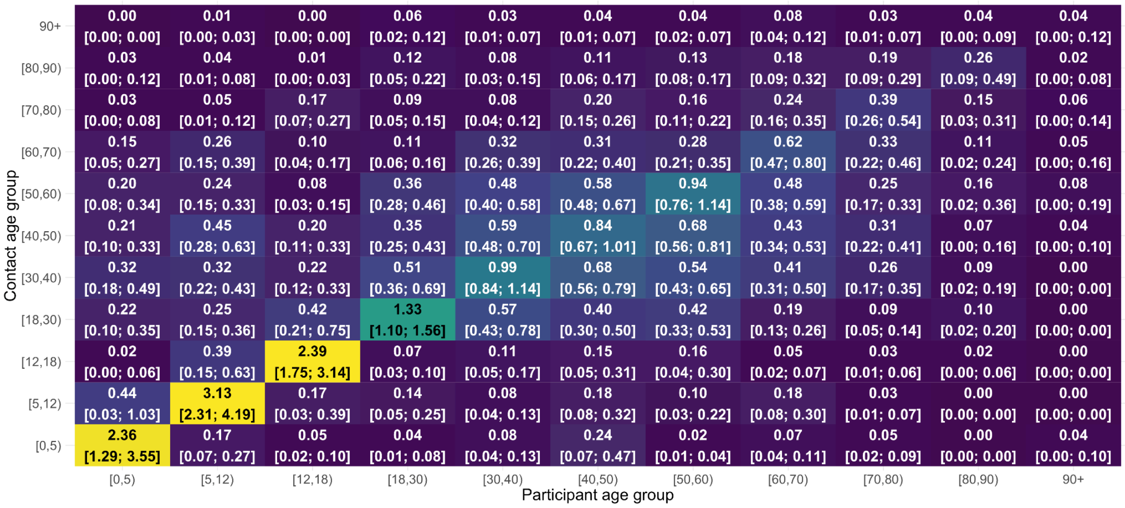

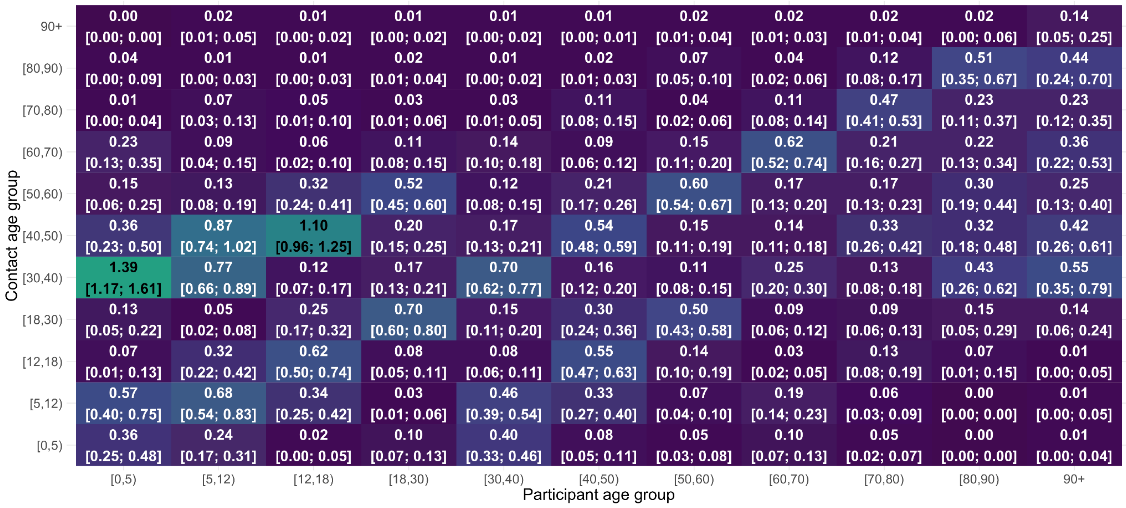


**Figure S4:** Social contact matrices for non-chronic participants *(Top)* outside the household *(Bottom)* inside the household contacts.

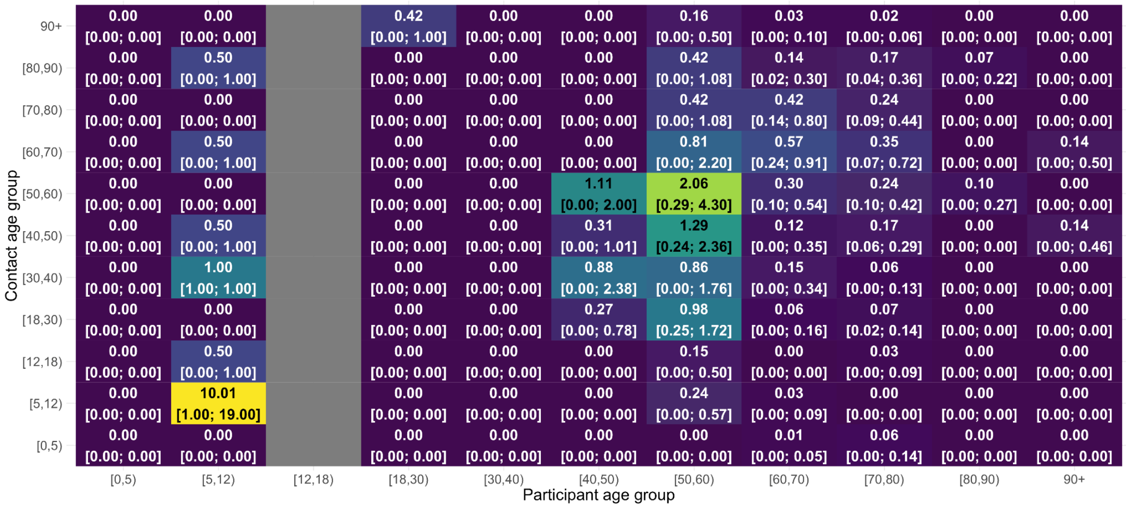


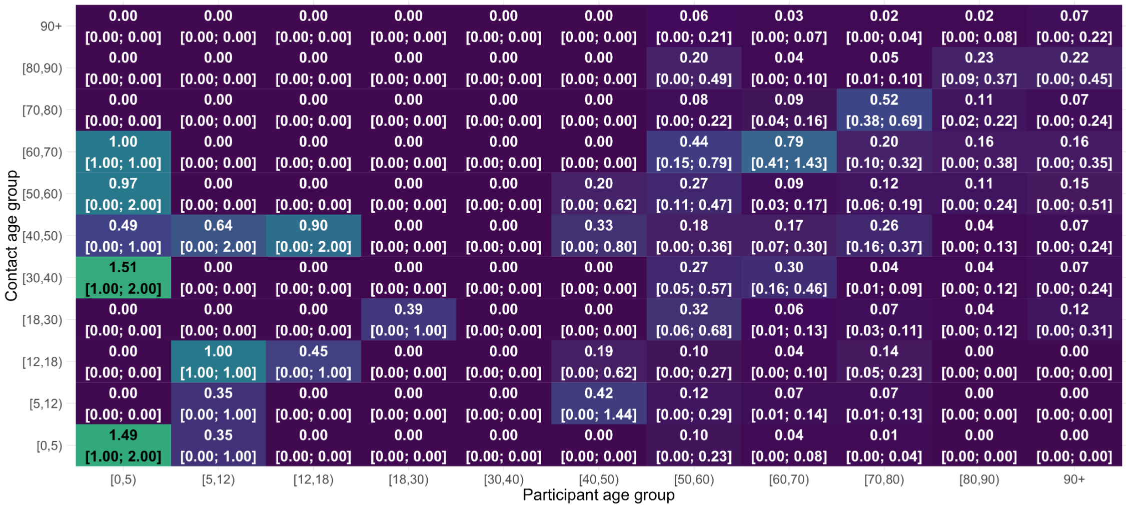


**Figure S5:** Social contact matrices for participants reporting no answer regarding their frailty level *(Top)* outside the household *(Bottom)* inside the household contacts.

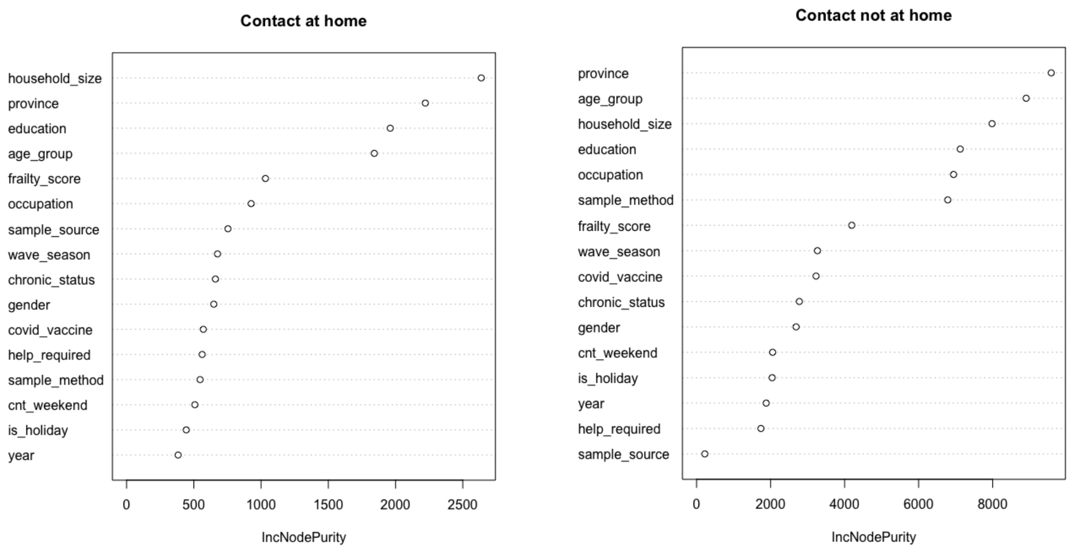


**Figure S6:** Variable importance calculated by the random forest

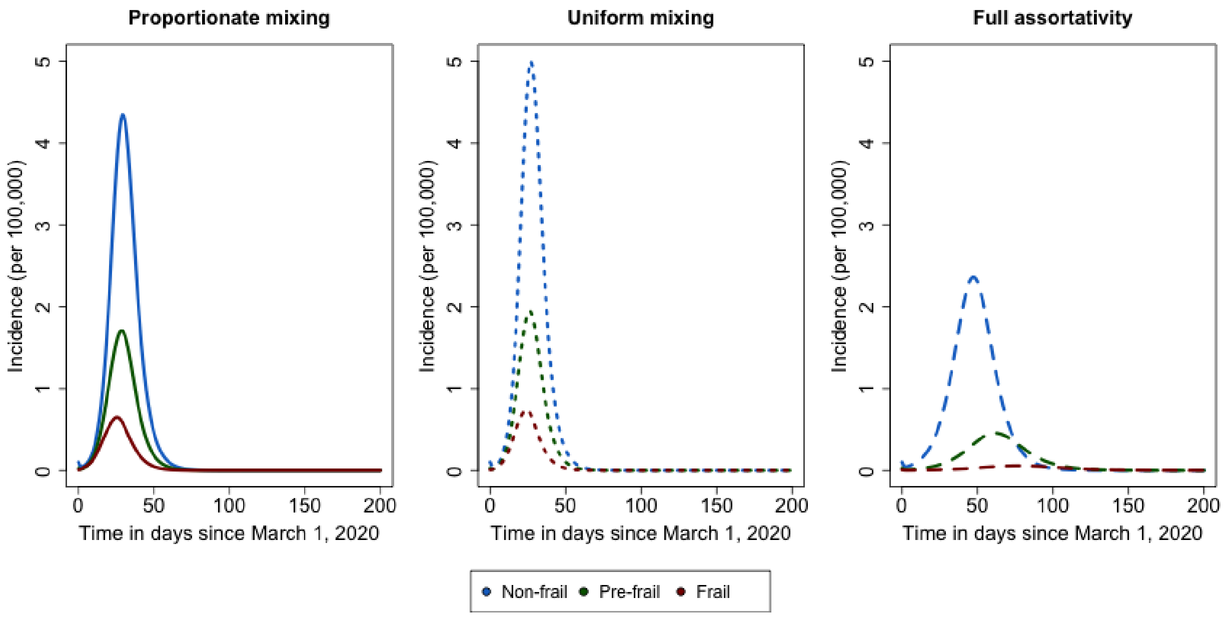


**Figure S7:** Comparison of COVID-19-like epidemic curves for the Belgian population with various frailty-based mixing patterns with the host- and disease-specific proportionality factor ($q$).

### 4. Supplementary materials (Tables)

| Study population | **General population** | **Care facilities** | **Chronic conditions** | **ILI** | **App** |
|---|---|---|---|---|---|
| Survey methods | Paper/Online | Face-to-face interview | Paper/Online | Paper/Online | App |
| Recruitment | National | List of | National | National | National |

| | | | | | |
|---|---|---|---|---|---|
| | Registry | facilities | Registry | Registry | Registry |
| Quota (Total) | 2097<br>(2529 + 1853) | 270 (271) | 170 (1160) | 250 (16) | 160 (166) |
| Total per age | 0-2: 143%<br>(325)<br>3-8: 118%<br>(177)<br>9-24: 113%<br>(373)<br>25-49: 96%<br>(458)<br>50-79: 124%<br>(933)<br>80-89: 142%<br>(185)<br>90-99: 130%<br>(78)<br><br>Boost sample:<br>21-99: 1853 | 50-75: 21%<br>(31)<br>75+: 198%<br>(238)<br><br>NA: 2 | 50-79: 717%<br>(1076)<br>80-89: 410%<br>(41)<br>90-99: 430%<br>(43) | 26-75: 3%<br>(16) | 21-60: 104%<br>(166) |

**Table S1:** Overview of study populations, survey methods, and quota in the data collection.

| Notation | Description | Value |
|---|---|---|
| $\gamma$ | Average length of latency period | 1.5 days |
| $\theta$ | Average length of pre-symptomatic infectious period | 2.9 days |
| $\delta_1$ | Average length of infectious period when mildly infected (after pre-symptomatic phase) | 2.4 days |
| $\delta_2$ | Average length of infectious period when mildly infected (after pre-symptomatic phase) | 6.5 days |
| $\delta_3$ | Recovery period of hospitalised individuals' length of stay in the hospital | 23.3 days |
| $\delta_4$ | Recovery period of individual length of stay in the ICU | 23.2 days |
| $q$ | Proportionality factor capturing contextual and host- and disease-specific characteristics | 0.051 |
| $\boldsymbol{\omega}$ | The average time between symptoms onset and hospitalisation. | 1.67 days |
| $\boldsymbol{\Phi}_1$ | The proportion of symptomatic cases developing only mild symptoms | 0.881 |
| $\boldsymbol{p}$ | The proportion of asymptomatic cases | 0.129 |

| | | |
|---|---|---|
| μ | The probability of dying upon hospitalisation (Number of deaths relative to number of hospitalised individuals) | 0.473 |

**Table S2:** Parameters utilised for the discrete-time age-structured SEIR compartmental model.

| Age group | Incidence | Age group | Incidence |
|---|---|---|---|
| 0-50 | 11,716 | 70-80 | 372 |
| 50-60 | 1,119 | 80-90 | 264 |
| 60-70 | 466 | 90-100 | 101 |

**Table S3:** Distribution of Initial COVID-19 Exposures Across Age Groups (March 1, 2020)

| | **Wave 1 (N = 535)** | **Wave 2 (N = 1,285)** | **Wave 3 (N = 3,388)** | **No answer (N = 515)** |
|---|---|---|---|---|
| **Survey date** | **2022-06-14 to 2023-06-26** | **2022-09-07 to 2023-01-20** | **2023-01-23 to 2023-05-09** | **-** |
| Unknown | 0 | 0 | 0 | 515 |
| **Sample source** | | | | |
| Nat rep | 466 (87%) | 1,174 (91%) | 3,298 (97%) | 514 (100%) |
| WZC | 69 (13%) | 111 (8.6%) | 90 (2.7%) | 1 (0.2%) |
| **Sample method** | | | | |
| Paper | 310 (58%) | 810 (63%) | 2,090 (62%) | 515 (100%) |
| CAWI | 225 (42%) | 467 (36%) | 1,140 (34%) | 0 (0%) |
| App | 0 (0%) | 8 (0.6%) | 158 (4.7%) | 0 (0%) |
| **Help required** | | | | |
| Self answered | 369 (69%) | 971 (76%) | 2,836 (84%) | 307 (60%) |
| With help | 159 (30%) | 294 (23%) | 486 (14%) | 61 (12%) |
| No answer | 7 (1.3%) | 20 (1.6%) | 66 (1.9%) | 147 (29%) |
| **Education** | | | | |
| Undergraduate | 97 (18%) | 291 (23%) | 967 (29%) | 51 (9.9%) |

| | | | | |
|---|---|---|---|---|
| degree | | | | |
| Diploma (primary education) | 61 (11%) | 139 (11%) | 267 (7.9%) | 61 (12%) |
| Diploma (secondary education) | 115 (21%) | 334 (26%) | 937 (28%) | 102 (20%) |
| Certificate (secondary education) | 39 (7.3%) | 92 (7.2%) | 256 (7.6%) | 46 (8.9%) |
| Postgraduate degree | 61 (11%) | 160 (12%) | 532 (16%) | 40 (5.8%) |
| No official diploma | 152 (28%) | 249 (19%) | 378 (11%) | 95 (18%) |
| No answer | 10 (1.9%) | 20 (1.6%) | 51 (1.5%) | 130 (2.5%) |
| **Province** | | | | |
| Antwerpen | 118 (22%) | 367 (29%) | 938 (28%) | 95 (18%) |
| Limburg | 80 (15%) | 165 (13%) | 517 (15%) | 68 (13%) |
| Oost-Vlaanderen | 135 (25%) | 265 (21%) | 695 (21%) | 81 (16%) |
| West-Vlaanderen | 106 (20%) | 209 (16%) | 574 (17%) | 97 (19%) |
| Vlaams Brabant | 82 (15%) | 226 (18%) | 544 (16%) | 50 (9.7%) |
| Others | 0 (0%) | 8 (0.6%) | 14 (0.4%) | 3 (0.6%) |
| No answer | 14 (2.6%) | 45 (3.5%) | 106 (3.1%) | 121 (23%) |
| **Gender** | | | | |
| Male | 220 (41%) | 597 (46%) | 1,503 (44%) | 208 (40%) |
| Female | 311 (58%) | 682 (53%) | 1,858 (55%) | 209 (41%) |
| Others | 0 (0%) | 0 (0%) | 2 (< 0.1%) | 0 (0%) |
| No answer | 4 (0.7%) | 6 (0.5%) | 25 (0.7%) | 98 (19%) |
| **Occupation** | | | | |
| Full time | 122 (23%) | 340 (26%) | 1,011 (30%) | 64 (12%) |
| Part time | 37 (6.9%) | 115 (8.9%) | 416 (12%) | 26 (5%) |
| Student job | 15 (2.8%) | 23 (1.8%) | 49 (1.4%) | 4 (0.8%) |
| Volunteer | 7 (1.3%) | 25 (1.9%) | 83 (2.4%) | 10 (1.9%) |

| | | | | |
|---|---|---|---|---|
| Unemployed | 221 (41%) | 560 (44%) | 1,524 (45%) | 274 (53%) |
| Not applicable | 128 (24%) | 201 (16%) | 263 (7.8%) | 24 (4.7%) |
| No answer | 5 (0.9%) | 21 (1.6%) | 42 (1.2%) | 113 (22%) |
| **Age group** | | | | |
| 0-9 | 109 (20%) | 154 (12%) | 174 (5.1%) | 16 (3.1%) |
| 10-19 | 31 (5.8%) | 73 (5.7%) | 132 (3.9%) | 16 (3.1%) |
| 20-29 | 39 (7.3%) | 74 (5.8%) | 212 (6.3%) | 8 (1.6%) |
| 30-39 | 43 (8%) | 93 (7.2%) | 315 (9.3%) | 16 (3.1%) |
| 40-49 | 40 (7.5%) | 93 (7.2%) | 302 (8.9%) | 20 (3.9%) |
| 50-59 | 42 (7.9%) | 190 (15%) | 562 (17%) | 38 (7.4%) |
| 60-69 | 68 (13%) | 242 (19%) | 818 (24%) | 95 (18%) |
| 70-79 | 60 (11%) | 181 (14%) | 610 (18%) | 112 (22%) |
| 80-90 | 59 (11%) | 108 (8.4%) | 120 (3.5%) | 47 (9.1%) |
| 90-100 | 39 (7.3%) | 71 (5.5%) | 102 (3.0%) | 30 (5.8%) |
| No answer | 5 (0.9%) | 6 (0.5%) | 41 (1.2%) | 117 (23%) |
| **Chronic status** | | | | |
| Chronic condition | 155 (29%) | 440 (34%) | 1,134 (33%) | 183 (36%) |
| No chronic condition | 285 (53%) | 730 (57%) | 2,024 (60%) | 162 (31%) |
| No answer | 95 (18%) | 115 (8.9%) | 230 (6.8%) | 170 (33%) |
| **Resident status** | | | | |
| Residential unit | 311 (58%) | 925 (72%) | 2,852 (84%) | 362 (70%) |
| With parents | 137 (26%) | 218 (17%) | 373 (11%) | 28 (5.4%) |
| Family other than parents | 4 (0.7%) | 7 (0.5%) | 15 (0.4%) | 0 (0%) |
| ROB | 19 (3.6%) | 24 (1.9%) | 43 (1.3%) | 1 (0.2%) |
| RVT | 27 (5%) | 64 (5%) | 19 (0.6%) | 1 (0.2%) |
| Others | 27 (5%) | 24 (1.9%) | 31 (0.9%) | 4 (0.8%) |
| No answer | 10 (1.9%) | 23 (1.8%) | 55 (1.6%) | 119 (23%) |
| **Household size** | | | | |

| | | | | |
|---|---|---|---|---|
| 1 | 42 (7.9%) | 124 (9.6%) | 392 (12%) | 54 (10%) |
| 2 | 137 (26%) | 399 (31%) | 1,224 (36%) | 139 (27%) |
| 3 | 83 (16%) | 216 (17%) | 675 (20%) | 76 (15%) |
| 4 | 129 (24%) | 232 (18%) | 540 (16%) | 28 (5.4%) |
| 5+ | 53 (9.9%) | 158 (12%) | 317 (9.4%) | 37 (7.2%) |
| ROB/RVT | 46 (8.6%) | 88 (6.8%) | 62 (1.8%) | 2 (0.4%) |
| No answer | 45 (8.4%) | 68 (5.3%) | 178 (5.3%) | 179 (35%) |
| **Holiday** | | | | |
| Yes | 279 (52%) | 122 (9.5%) | 541 (16%) | 0 (0%) |
| No | 256 (48%) | 1,163 (91%) | 2,847 (84%) | 0 (0%) |
| No answer | 0 (0%) | 0 (0%) | 0 (0%) | 515 (100%) |
| **Year** | | | | |
| 2022 | 360 (67%) | 1,251 (97%) | 0 (0%) | 0 (0%) |
| 2023 | 175 (33%) | 34 (2.6%) | 3,388 (100%) | 0 (0%) |
| No answer | 0 (0%) | 0 (0%) | 0 (0%) | 515 (100%) |
| **Frailty score** | | | | |
| Frail | 90 (17%) | 204 (16%) | 448 (13%) | 88 (17%) |
| Pre-frail | 129 (24%) | 366 (28%) | 1,153 (34%) | 101 (20%) |
| Non-frail | 293 (55%) | 651 (51%) | 1,617 (48%) | 120 (23%) |
| Missing | 23 (4.3%) | 64 (5%) | 170 (5%) | 206 (40%) |

**Table S4:** Summary statistics of the Epicurus study. Summary statistics for numeric variables are median (IQR) values and any unanswered questions (marked as NA or "No answer") in the survey were excluded from the table. Consequently, the percentages displayed in the table will not sum up to 100%.

| | **Wave 1 (N = 3,110)** | **Wave 2 (N = 7,218)** | **Wave 3 (N = 19, 392)** | **No answer (N = 1,655)** |
|---|---|---|---|---|
| **Contact date**<br><br>Unknown | **2022-06-13 to 2023-06-25**<br><br>**0** | **2022-09-06 to 2023-01-19**<br><br>**0** | **2023-01-22 to 2023-05-08**<br><br>**0** | **-**<br><br>**1,655** |
| **Type of day (contact)** | | | | |

| | | | | |
|---|---|---|---|---|
| Weekday | 2,475 (80%) | 5,670 (79%) | 15,312 (79%) | 0 (0%) |
| Weekend | 645 (20%) | 1,548 (21%) | 4,080 (21%) | 0 (0%) |
| No answer | 0 (0%) | 0 (0%) | 0 (0%) | 1,655 (100%) |
| **Contact at home** | 1,393 (45%) | 3,043 (42%) | 7,348 (38%) | 522 (32%) |
| **Physical contact** | | | | |
| Yes | 1,806 (58%) | 3,799 (53%) | 9,121 (47%) | 644 (39%) |
| No | 918 (30%) | 3,001 (42%) | 7,742 (40%) | 401 (24%) |
| No answer | 386 (12%) | 418 (5.8%) | 2,529 (13%) | 610 (37%) |
| **Contact duration** | | | | |
| ≤ 5 minutes | 152 (4.9%) | 372 (5.2%) | 954 (4.9%) | 66 (40%) |
| 5 - 15 minutes | 371 (12%) | 763 (11%) | 1,815 (9.4%) | 90 (5.4%) |
| 15 min - 1 hour | 389 (13%) | 1,277 (18%) | 3,244 (17%) | 190 (11%) |
| 1 - 4 hours | 741 (24%) | 2,041 (28%) | 5,620 (29%) | 360 (22%) |
| ≥ 4 hours | 1,068 (34%) | 2,416 (33%) | 5,437 (28%) | 438 (26%) |
| No answer | 389 (13%) | 349 (4,8%) | 2,322 (12%) | 521 (31%) |
| **Number of contacts** | 5 (2, 7) | 4 (2, 7) | 5 (3, 7) | 1 (0, 4) |

**Table S5:** Summary statistics of the reported contacts (Median (IQR))

| | **p-value** | |
|---|---|---|
| | **Contacts at home** | **Contacts not at home** |
| Sample source | < 0.05 | 0.247 |
| Help required | < 0.05 | 0.256 |
| Chronic status | 0.445 | 0.147 |
| Covid-19 vaccination status | 0.977 | 0.139 |
| Education | 0.982 | < 0.05 |
| Province | 0.193 | 0.401 |
| Age group: Frailty score | 0.348 | 0.477 |

| | | |
|---|---|---|
| Holiday: Contact day | 0.851 | < 0.05 |
| Age group: Contact day | 0.712 | 0.249 |
| Age group: Holiday | 0.133 | < 0.05 |
| Frailty score: Help required | < 0.05 | 0.487 |

**Table S6:** The likelihood ratio test for GAMLSS, p-value <0.05 indicates that the variable is retained or added to the model. If a variable is significant in either model (at home or not at home), it will be used in both GAMLSS models.

| Covariates | RI (Outside) | RI (Inside) |
|---|---|---|
| **Sample source** | | |
| General | | |
| Care facilities | 0.905 [0.681; 1.202] | 2.141* [1.797; 2.550] |
| **Help required** | | |
| Self answered | | |
| With help | 0.825 [0.662; 1.028] | 1.497* [1.325; 1.692] |
| No answer | 1.229 [0.723; 2.087] | 0.929 [0.623; 1.386] |
| **Age group** | | |
| 0-9 | - | - |
| 10-19 | 1.345 [0.905; 1.999] | 1.258* [1.033; 1.532] |
| 20-29 | 0.960 [0.596; 1.547] | 1.089 [0.827; 1.435] |
| 30-39 | 0.855 [0.539; 1.357] | 1.063 [0.799; 1.413] |
| 40-49 | 0.605* [0.373; 0.981] | 1.152 [0.870; 1.527] |
| 50-59 | 0.679 [0.433; 1.063] | 1.065 [0.816; 1.390] |
| 60-69 | 0.671 [0.434; 1.036] | 1.173 [0.907; 1.518] |
| 70-79 | 0.616* [0.395; 0.962] | 1.210 [0.928; 1.577] |
| 80-89 | 0.613 [0.351; 1.072] | 1.163 [0.820; 1.649] |
| 90-100 | 0.599 [0.330; 1.085] | 0.847 [0.549; 1.309] |
| No answer | 0.944 [0.458; 1.947] | 1.174 [0.662; 2.083] |
| **Gender** | | |

| | | |
|---|---|---|
| Male | - | - |
| Female | 1.077* [1.012; 1.146] | 1.066* [1.020; 1.113] |
| Others | 1.074 [0.278; 4.144] | 2.676* [1.202; 5.956] |
| No answer | 0.680 [0.456; 1.012] | 1.153 [0.862; 1.541] |
| **Household size** | | |
| 1 | - | - |
| 2 | 0.854* [0.770; 0.948] | 1.233* [1.111; 1.367] |
| 3 | 0.859* [0.764; 0.964] | 1.562* [1.403; 1.740] |
| 4 | 0.882 [0.776; 1.003] | 2.009* [1.799; 2.242] |
| 5+ | 0.866* [0.753; 0.995] | 2.315* [2.060; 2.602] |
| ROB/RVT | 0.733 [0.529; 1.014] | 1.249* [1.034; 1.510] |
| No answer | 0.835* [0.718; 0.972] | 1.269* [1.103; 1.460] |
| **Sample method** | | |
| Paper | - | - |
| CAWI | 0.436* [0.406; 0.468] | 0.862* [0.823; 0.904] |
| App | 0.404* [0.337; 0.485] | 0.886 [0.782; 1.005] |
| **Occupation** | | |
| Full time | - | - |
| Part time | 0.914 [0.824; 1.014] | 0.987 [0.913; 1.066] |
| Student job | 0.719* [0.543; 0.950] | 0.927 [0.780; 1.102] |
| Volunteer | 1.010 [0.817; 1.250] | 1.194* [1.012; 1.409] |
| Unemployed | 0.641* [0.573; 0.716] | 1.087* [1.001; 1.180] |
| Not applicable | 0.800 [0.565; 1.133] | 1.158 [0.953; 1.407] |
| Not answer | 0.829 [0.636; 1.080] | 1.219 [0.987; 1.507] |
| **Frailty score** | | |
| Frail | - | - |
| Pre-frail | 1.095 [0.963; 1.246] | 0.992 [0.906; 1.086] |
| Non-frail | 1.175* [1.029; 1.342] | 1.023 [0.932; 1.123] |
| Missing | 0.900 [0.751; 1.077] | 0.899 [0.777; 1.041] |

| | | |
|---|---|---|
| **Education** | | |
| Undergraduate degree | - | - |
| Diploma (primary education) | 0.599* [0.513; 0.699] | 0.978 [0.888; 1.078] |
| Diploma (secondary education) | 0.778* [0.717; 0.843] | 1.002 [0.942; 1.065] |
| Certificate (secondary education) | 0.709* [0.617; 0.814] | 0.981 [0.890; 1.081] |
| Postgraduate degree | 1.052 [0.959; 1.154] | 1.005 [0.937; 1.078] |
| No official diploma | 0.703* [0.603; 0.821] | 1.021 [0.922; 1.131] |
| No answer | 0.724* [0.571; 0.918] | 0.948 [0.759; 1.185] |
| **Year: Wave** | | |
| 2022: Wave 1 (Summer) | 0.878 [0.771; 1.000] | 0.930 [0.849; 1.017] |
| 2023: Wave 1 (Summer) | 1.154 [0.957; 1.391] | 1.097 [0.986; 1.220] |
| 2022: Wave 2 (Fall) | 0.870* [0.808; 0.937] | 0.993 [0.944; 1.045] |
| 2023: Wave 2 (Fall) | 0.850 [0.586; 1.232] | 0.941 [0.732; 1.209] |
| 2023: Wave 3 (Winter) | - | - |
| **Holiday: Day of contact** | | |
| Holiday (No): Weekend | - | - |
| Holiday (No): Weekday | 1.319* [1.218; 1.427] | 0.854* [0.810; 0.901] |
| Holiday (Yes): Weekend | 0.791 [0.592; 1.057] | 0.963 [0.817; 1.135] |
| Holiday (Yes): Weekday | 0.780 [0.606; 1.005] | 0.818* [0.709; 0.943] |
| **Age group: Holiday** | | |
| 0-9: Holiday (No) | - | - |
| 10-19: Holiday (No) | 0.801 [0.531; 1.209] | 0.749* [0.608; 0.922] |
| 20-29: Holiday (No) | 0.577* [0.394; 0.847] | 0.874 [0.696; 1.098] |
| 30-39: Holiday (No) | 0.574* [0.409; 0.804] | 1.004 [0.800; 1.260] |
| 40-49: Holiday (No) | 0.907 [0.626; 1.314] | 0.916 [0.736; 1.142] |
| 50-59: Holiday (No) | 0.729* [0.535; 0.995] | 0.905 [0.743; 1.103] |
| 60-69: Holiday (No) | 0.725* [0.541; 0.971] | 0.849 [0.702; 1.026] |
| 70-79: Holiday (No) | 0.662* [0.485; 0.904] | 0.819* [0.670; 1.000] |
| 80-89: Holiday (No) | 0.554* [0.345; 0.888] | 0.769 [0.564; 1.048] |

| | | |
|---|---|---|
| 90-100: Holiday (No) | 0.495* [0.292; 0.839] | 1.352 [0.902; 2.027] |
| No answer: Holiday (No) | 0.503 [0.251; 1.007] | 0.843 [0.467; 1.519] |
| **Frailty score: Help required** | | |
| Non-frail: With help | 1.095 [0.822; 1.458] | 0.685* [0.584; 0.803] |
| Pre-frail: With help | 1.218 [0.907; 1.635] | 0.779* [0.654; 0.927] |
| Missing: With help | 1.005 [0.584; 1.728] | 0.739 [0.508; 1.075] |
| Frail: Self answered | - | - |
| Non-frail: No answer | 0.846 [0.458; 1.563] | 1.324 [0.840; 2.087] |
| Pre-frail: No answer | 0.588 [0.308; 1.121] | 0.903 [0.525; 1.551] |
| Missing: No answer | 0.669 [0.319; 1.405] | 0.841 [0.413; 1.714] |

**Table S7:** NBI Generalised Linear Model Summary Statistics (95% CI) for Outside- and Inside- home contacts. The asterisk (*) indicates significance of the variable (p-value < 0.05)
